# The Effects of Heart Rate Variability Biofeedback on Functional Gastrointestinal Disorders: A Scoping Review

**DOI:** 10.1101/2024.07.22.24310788

**Authors:** Ashley G Pereira, Lily Fu, William Xu, Armen A Gharibans, Greg O’Grady

**Author notes:** **Corresponding author** Prof. Greg O’Grady, Department of Surgery University of Auckland Auckland, New Zealand.

## Abstract

Functional Gastrointestinal Disorders (FGID) are a group of symptom-based disorders that occur across the alimentary tract and have a high prevalence globally in both adults and children. These symptoms are chronic and/or recurrent and often have substantial effects on quality of life. Their incidence is tied to multiple factors, including gut-brain axis imbalance, which includes autonomic dysregulation related to a relative withdrawal of vagal activity. Heart rate variability biofeedback (HRVB) is a non-invasive intervention that can influence autonomic activity and has shown benefit for diverse conditions including depression and anxiety, however the evidence of its effect has not yet been systematically assessed in FGIDs. This scoping review aimed to collate and evaluate the available literature regarding HRVB and FGIDs. We systematically searched four medical databases. Four articles met inclusion criteria for being interventional studies using HRVB in FGIDs. These were heterogeneous, including both paediatric and adult as well different subtypes of FGID. Two of the four studies demonstrated significant improvements from HRVB interventions in FGID symptoms while the other two found no significant difference. Scoping evaluation indicated this inconsistency likely reflects heterogeneous populations and study designs. Further scoping review of the broader HRVB literature also discovered that at least six weeks of HRVB is required to observe an impact on FGID symptoms and defined recommended guidance for performing future evaluations of HRVB in FGIDs. Evidence on HRVB for FGID is emergent, however HRVB appears a promising intervention when administered optimally. Further studies using best-practice techniques are required.

## Introduction

Functional Gastrointestinal Disorders (FGID), more recently termed Disorders of Gut-Brain Interactions (DGBIs), are a group of multiple symptom phenotypes that occur across the gastrointestinal (GI) tract. There are several subtypes and symptoms range from dysphagia to dyspepsia to abdominal pain and bloating (Black et al. 2020). These disorders may have recurrent and potentially debilitating impacts, and an incomplete understanding of their pathophysiology means that clinical diagnosis and treatment often still rely upon trial and error. They are highly prevalent, affecting up to 40% of the global population (Black et al. 2020; Sperber et al. 2021); (Andreasson et al. 2021).

Recently, there has been increasing evidence of a correlation between the prevalence of functional gut symptoms and an imbalance of autonomic nervous system activity, with greater relative sympathetic activity due to parasympathetic withdrawal (Aggarwal et al. 1994; Bharucha et al. 1993; Mróz, Czub, and Brytek-Matera 2022); (Jung-Ho et al. 1999). This hypothesis is supported by the emerging efficacy of therapies proposed to enhance vagal tone, encompassing such diverse approaches as chewing gum, slow breathing exercises, moderate-pressure massage, or transcutaneous vagal electrical stimulation (Lunding et al. 2008; Zhang et al. 2023), accompanied by with evidence for improved antral, colonic and oesophageal motility and symptom reductions (Bonaz, Sinniger, and Pellissier 2016).

Heart Rate Variability Biofeedback (HRVB) is a non-invasive technique that leverages heart rate variability and autonomic regulation principles, using specific breathing rates to modulate heart rate, enhance baroreflex sensitivity, and balance the autonomic nervous system by increasing relative parasympathetic drive (P. M. Lehrer and Gevirtz 2014). Regular practice of HRVB can also promote neuroplasticity and improve the adaptability and flexibility of the vagal system, increasing baroreflex gain while at rest (Wheat and Larkin 2010). This ability to influence the activity of the autonomic nervous system could therefore have clinical significance for FGIDs. Studies have been conducted to investigate this potential relationship. However, to our knowledge there has been no systematic study collating the evidence for HRVB on the GI tract. In addition, standardised protocols for performing HRVB have not been implemented; although the most common protocol is that of Lehrer et al (P. Lehrer et al. 2013), there still exists considerable variation.

The aim of this study was therefore to examine the role and effectiveness of HRVB as a potential therapy for FGIDs through a scoping review. The primary aim was to identify and assess relevant clinical studies applying HRVB as an intervention to reduce FGID symptoms. The secondary aims were to assess the protocols and measurement tools used by each study, in order to develop a protocol for future studies to measure the effect of biofeedback on patients diagnosed with FGIDs, and to guide future research in this emerging area.

## Methods

### Study Design

The scoping review was conducted and reported in accordance with the PRISMA 2020 guidelines and the scoping review extension (Tricco et al. 2018; Page et al. 2021).

### Search Strategy and Study Selection

Four databases were searched: PubMed, Web of Science, ScienceDirect and Scopus; with ‘Heart Rate Variability’ alongside different categories and terminologies for FGIDs including “Functional abdominal pain”, “Nausea and Vomiting Syndromes”, “Functional Dyspepsia”, “Gastroparesis”, “Irritable Bowel Syndrome”, “Functional Constipation”, “Functional Diarrhoea”, and “Functional abdominal bloating” (i.e. “Heart Rate Variability AND Functional Dyspepsia). The literature search was completed on 8th January 2024.

It was decided to use the search term ‘heart rate variability’ as opposed to ‘heart rate variability biofeedback’ as it was a broader search term, and many studies did not use this term in their work, instead opting for ‘slow deep breathing’ or similar phrases to describe the same technique.

Two reviewers independently screened the literature titles, abstracts and then the entire article according to the inclusion and exclusion criteria detailed in **Table 1**. There were no limits on the method by which HRV or gastrointestinal symptoms were measured. Studies that used ECG and PPG. were both collected as both were proved to be equivalent to each other as justified by Plews et al. (Plews et al. 2017), as well as studies that used both symptom questionnaires and concurrent clinical investigations.

**Table 1:** The key inclusion and exclusion criteria that was used to screen all relevant articles.

| <u>Inclusion Criteria</u> | <u>Exclusion Criteria</u> |
| --- | --- |
| Measures heart rate variability | Does not measure Heart Rate Variability |
| Intervention of HRVB or similar as per Lehrer et al. 2013 | Gastrointestinal syndromes of known physiological cause (non FGIDs) |
| Published within the last 10 years (2014 onwards) | Intervention is not solely heart rate variability biofeedback or auricular transcutaneous electrical stimulation |
|  | Articles whose clinical focus is outside the scope of the search |
|  | Observational Studies |
|  | Conference abstracts and review articles |
|  | Articles in languages other than English |

### HRV Metrics

To assess HRV, there are several different metrics that can be used, and the following considerations were incorporated into the review. HRV metrics are primarily divided into Time Domain and Frequency Domain Measures (Fred Shaffer and Ginsberg 2017). The Time Domains largely focus on the interbeat intervals (IBI) or the time between each successive heartbeat, displaying the variance in these successive intervals. The two most commonly used metrics for this are SDNN (Standard Deviation of the N to N Intervals) and RMSSD (Root Mean Square of the Successive Differences). There are other variables which are included within the time domain measurements which were not relevant to the scope of this review. The Frequency Domain Measures rely on the ability to conduct a Fast Fourier Transform (FFT) on the heart rate data, separating the data into three separate bands: high frequency, low frequency, and very low frequency (HF, LF and VLF, respectively). Each of these frequency bands cover a set range of frequencies: 0.15–0.40 Hz for HF, 0.04–0.15 Hz for the LF, and 0.0033–0.04 Hz for VLF, and they are expressed as a power within those frequency bands. The final metric that is sometimes used, which is simply a calculation, is the Baevsky Stress Index (SI) (refer to **Figure 2**). This is a geometric method to assess IBIs and represent the function of the sympathetic nervous system.

**Figure 1:**
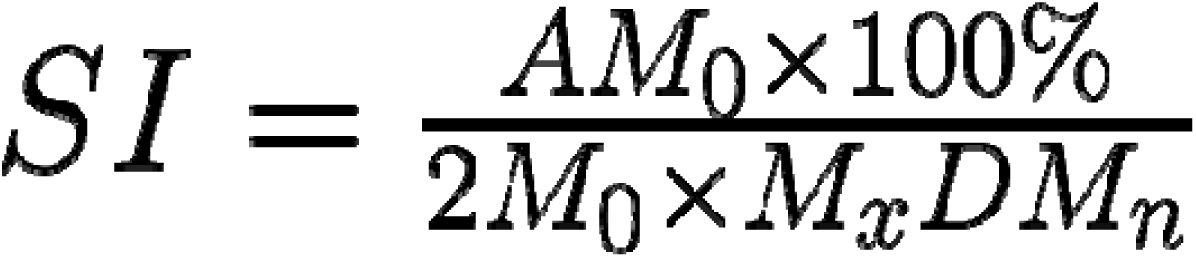
Baevsky Stress Index Calculation. M_0_ is the mode, AM_0_ is the mode amplitude calculated using a 50 ms bin width, M_x_DM_n_ is the difference between the longest (M_x_) and the shortest (M_n_) interval.

**Figure 2:**
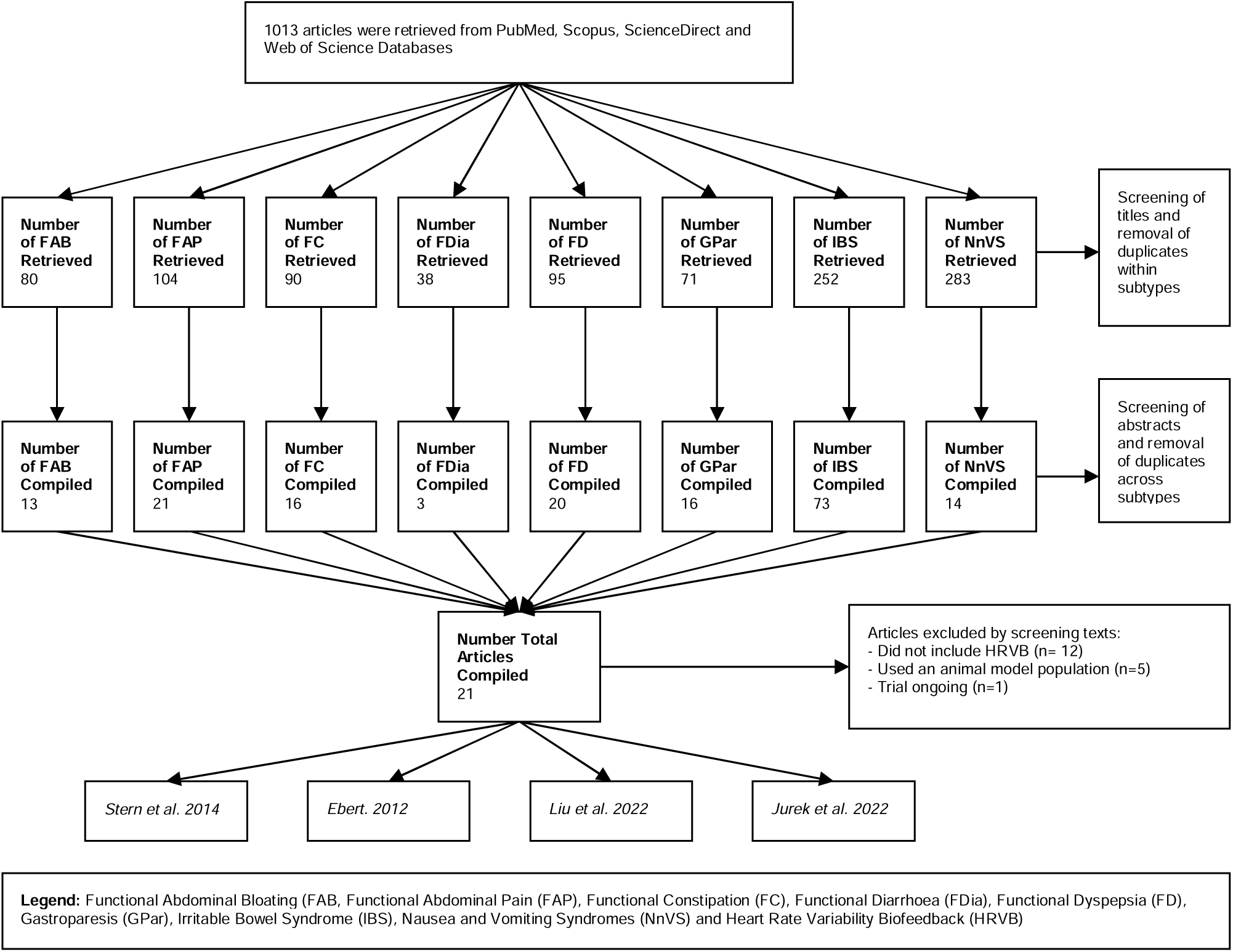
A diagramatic representation of the screening process of the articles retrieved for this review completed by both independent reviewers.

### Data Extraction and Analysis

Records from each database search were screened for inclusion by two independent authors, with discrepancies being discussed and resolved by mediation by a third author as required. Relevant data from the full-text articles were extracted independently then compared. Due to the heterogenous design as well as the limited number of studies available to be statistically combined, meta analysis was not performed. A narrative scoping review was therefore conducted on the included studies, allowing the reviewers to assess the role of HRV Biofeedback as a potential therapy for FGID and examine the protocols employed by each of the studies per the study aims in order to guide future research in this field.

## Results

### Literature Search Results

This literature search had resulted in a total of 1013 articles (including duplicates) with the following breakdown: 80 results for functional abdominal bloating, 104 results for functional abdominal pain, 90 for functional constipation, 38 for functional diarrhoea, 95 for functional dyspepsia, 71 for gastroparesis, 252 for irritable bowel syndrome and 283 for nausea and vomiting syndrome. The titles and abstracts of these articles were then screened independently by both reviewers according to the exclusion criteria as well as removing duplicates. This resulted in 4 total articles with some of them assessing multiple of the disorder subtypes that were included in the literature search (2 for functional abdominal pain, 1 for functional constipation, and 3 for irritable bowel syndrome). The data from these papers was then extracted and analysed. A graphical summary of the systematic literature review is presented in **Fig. 2**.

### Article Characteristics

From the four relevant studies identified in the literature search, three were conducted in the USA (Stern, Guiles, and Gevirtz 2014; Ebert 2012; Katherine Jurek et al. 2022) and one in China (Liu et al. 2022). The majority of these addressed IBS, while some studies addressing functional abdominal pain, functional constipation, as well as other subtypes, were found to be lacking (**Table 2**).

**Table 2:**
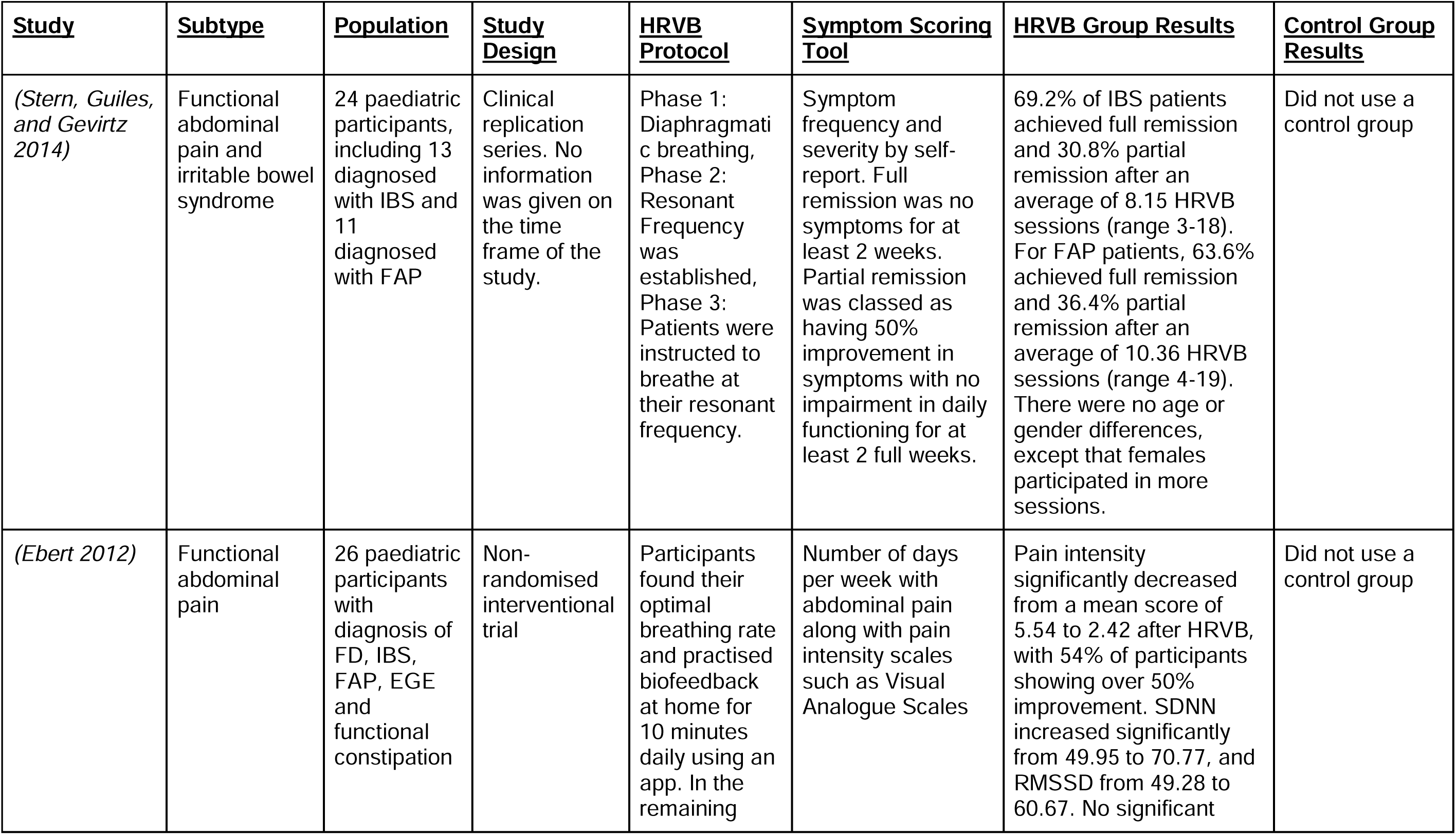
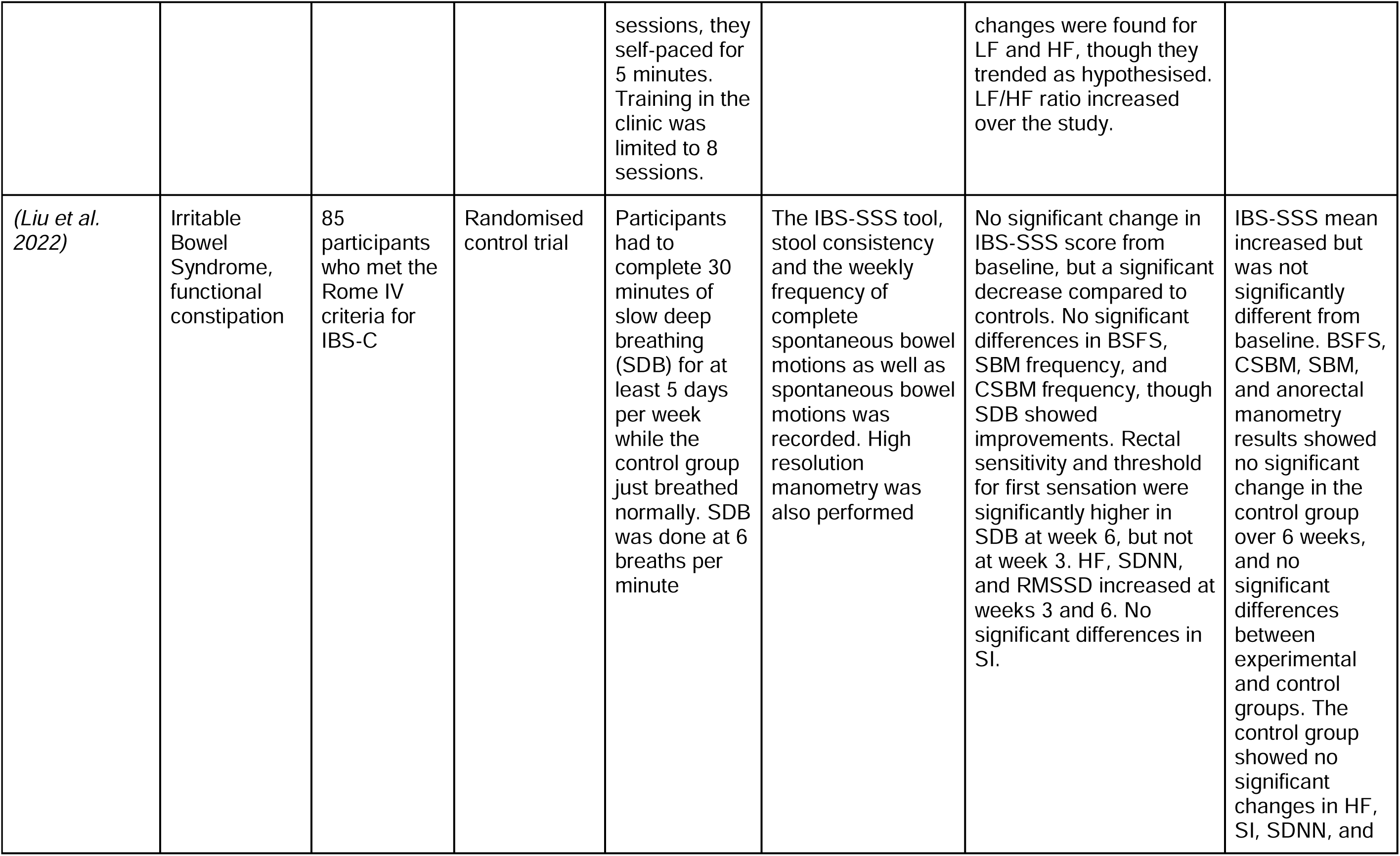
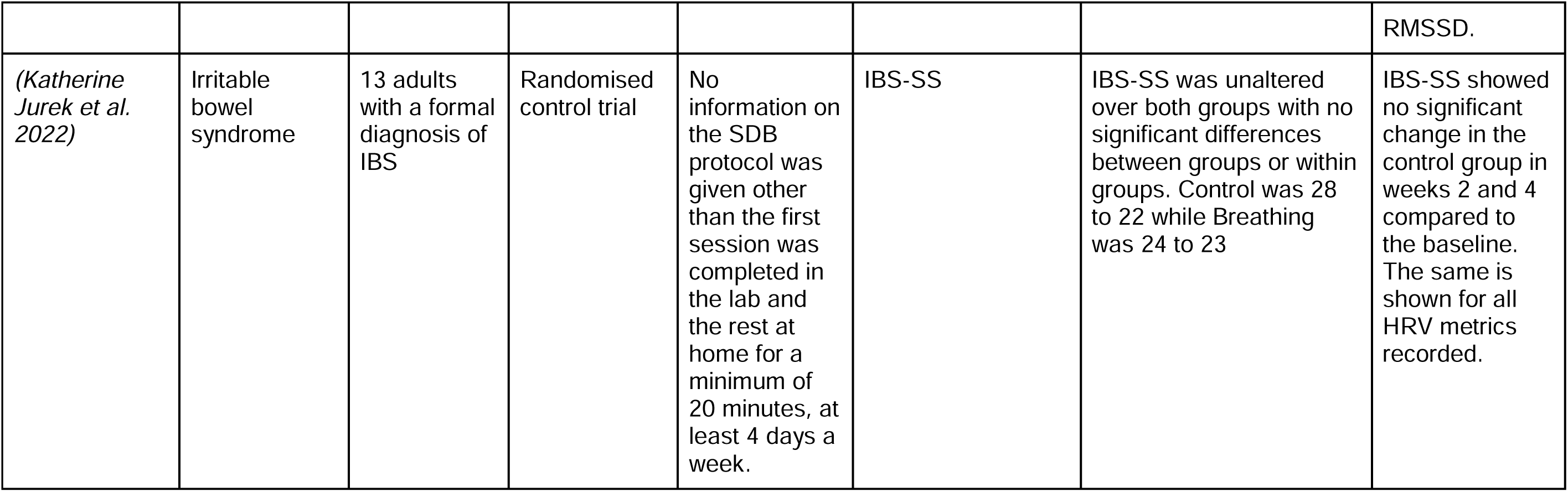
Summary of the key data from the four studies that resulted from the literature search and inclusion and exclusion criteria.

Two of the studies were randomised control trials (Liu et al. 2022; Katherine Jurek et al. 2022) while the other two were interventional studies where HRV biofeedback was not compared to a sham control scenario (Stern, Guiles, and Gevirtz 2014; Ebert 2012). These four articles also varied in terms of the target population, with two focused on paediatric populations (Stern, Guiles, and Gevirtz 2014; Ebert 2012) and the other two on adult populations (Liu et al. 2022; Katherine Jurek et al. 2022). (**Table 2**)

All of the included studies used the same primary metrics when quantifying the HRV present, relying on time and frequency domain metrics, to make inferences of vagal and sympathetic tone (Katherine Jurek et al. 2022). All four studies also used electrocardiograms as the method of measuring HRV data while participants were in the research laboratory/clinic. For the at-home biofeedback training, Stern et al used a portable, handheld device, which had photoplethysmography (PPG) capabilities called the StressEraser (Helicor Inc, New York, United States of America). Jurek et al opted to use a video to guide participants through their biofeedback at home which did not collect heart rate data. Liu et al and Ebert did not detail any at-home biofeedback practice. All of the four studies used different software to calculate and assess the HRV metrics stated above. Both Stern et al and Ebert used the J & J Engineering I-330 C-2+ hardware and Stern et al stated that they used the J & J Engineering USE3 software along with it, which is a combination of hardware and associated software to conduct biofeedback and measure HRV in the lab as well as calculate the metrics related to HRV (**Table 3**).

**Table 3:** Summary of the HRV metrics measured and used as part the respective analysis sections of each of the five studies.

| **<u>Study</u>** | **<u>Metrics Used</u>** | **<u>Method of Measurement</u>** | **<u>Method of Analysis</u>** |
| --- | --- | --- | --- |
| *(Stern, Guiles, and Gevirtz 2014)* | Heart Rate, Frequency domain spectral analysis | ECG | J & J Engineering I-330 C-2+ |
| *(Ebert 2012)* | SDNN, RMSSD, Frequency domain spectral analysis | ECG | J & J Engineering I-330 C-2+ Portable 6-Channel Physiological Monitoring System |
| *(Liu et al. 2022)* | Frequency domain spectral analysis, Stress Index, RMSSD and SDNN | ECG | Not stated |
| *(Katherine Jurek et al. 2022)* | RMSSD, LF/HF, Stress Index, SD2 | ECG (Polar Heart Rate Monitor) | Kubios |

The studies all used different techniques to measure and analyse HRV data, although common themes were identified. Stern et al and Ebert used similar technology, both using the previously mentioned J & J Engineering I-330 C-2+ system which has the ability to non-invasively measure heart rate via an ECG or PPG as well as respiration through a respiration belt. The system also has the ability to measure supplementary metrics of autonomic activity such as muscle tension, skin conductance and skin temperature, although these metrics were not analysed and reported within Stern et al’s trial but were in Ebert’s. Liu et al did not state what system they used to analyse HRV data. Jurek et al instead used a Polar Heart Rate Monitor (Polar Electro Oy, Kempele, Finland), which is a belt that goes across the chest with attached ECG electrodes. The raw ECG data from this was then fed through the Kubios software (Kubios Oy, Kuopio Finland), which has the ability to easily analyse HRV data as long as the operator can accurately identify and delete any ectopic beats from the data. Liu et al did not state how they analysed their HRV data from the ECG data they collected. None of the articles mentioned how they removed any potential artefact from their data as part of their analysis.

### HRV Biofeedback Protocols

All of these articles used HRV biofeedback as a form of intervention within their exposure groups. There is a considerable amount of variation present in the HRVB protocols being currently used, but the core structure is that it is initially started by providing some patient information to ensure patient buy-in, followed by equipment set-up, and an explanation of the physiological measures displayed on screen. After this, participants are guided through a biofeedback session of slow, controlled breathing while their heart rate variability is being simultaneously measured. This occurs for a set period of time, often at least 10 minutes while the participant attempts to maintain a mindful state, relaxing and focusing on their breathing. After the initial session, the participant then completes biofeedback training at home for a set period of time.

The main points of variation emerged when considering the breathing paces used for each study as well as the HRV Metrics used. The first point of variation was whether study investigators instruct the participants to breathe at a standardised breathing frequency (i.e. 6 breaths per min) or at the participants’ resonance frequency. (**Table 4**) Another point of variance in the protocol between the four included studies was the length of time during which they conducted the biofeedback training intervention. All four studies appeared to conduct studies of at least four weeks or more (excluding Stern et al and Jurek et al who did not state the timeframe of their study). This included time for at-home practice with a pacer device or a smartphone app that paced the individuals breathing along with a measurement of their heart rate via a PPG, averaging 120 minutes total per week (**Table 4**).

**Table 4:** Summary of the HRV Biofeedback and control protocols used as part the respective analysis sections of each of the five studies.

| <u>Study</u> | <u>Total Intervention Time</u> | <u>Implementation of Resonance Frequency</u> | <u>Breathing Frequency (breaths per min)</u> | <u>Total Minimum Minutes per Week of At-Home HRVB Training</u> | <u>Control Group Protocol</u> |
| --- | --- | --- | --- | --- | --- |
| *(Stern, Guiles, and Gevirtz 2014)* | Not stated | Yes | N/a | 140 minutes | None |
| *(Ebert 2012)* | Not stated (8 clinic sessions total) | Yes | N/a | 70 minutes | None |
| *(Liu et al. 2022)* | 6 weeks | No | 6 bpm | 150 minutes | Regular breathing |
| *(Katherine Jurek et al. 2022)* | 4 weeks | Not stated | Not stated | Not stated | Regular activities |

### Gastrointestinal Outcome Measures

The primary outcome measured across all four studies was a change in GI symptoms. The most common symptom scoring tool used was the IBS-SSS (used by Liu et al and Jurek et al), a validated questionnaire that assesses the severity of IBS according to four domains: pain intensity, frequency, location and relation to stool pattern. The remaining studies used symptom frequency and severity as common measures although there was no formal tool used other than a visual analogue scale. (**Table 5**)

**Table 5:** Summary of the gastrointestinal symptoms measured, and other clinical measures used as part the respective analysis sections of each of the four studies.

| <u>**Study**</u> | <u>**Symptom Measure**</u> | <u>**Other Clinical Measure**</u> |
| --- | --- | --- |
| *(Stern, Guiles, and Gevirtz 2014)* | Self-reported symptom severity and frequency as well as impairment of daily functioning | None |
| *(Ebert 2012)* | Number of reported days per week with abdominal pain along with pain intensity scales using a visual analogue scale | None |
| *(Liu et al. 2022)* | IBS-SSS, Stool consistency and weekly frequency of spontaneous bowel motions and complete spontaneous bowel motions. | High resolution anorectal manometry |
| *(Katherine Jurek et al. 2022)* | IBS-SS | None |

Out of the four studies, none used multiple physiological outcome measures. Liu et al was the only study to include a singular physiological outcome measure of high resolution anorectal manometry.

### Study Outcomes

Only two of the studies assessed in-depth as part of the scoping review, showed a significant improvement in patient outcomes. Stern et al and Liu et al were able to display evidence of the beneficial effects of biofeedback as an emerging therapy. There was a statistically significant decrease in symptom severity and frequency after the biofeedback trial had been completed, compared to baseline; with Stern demonstrating complete remission in 69.2% of participants and Liu et al. showing a statistically significant improvement in IBS-SSS and stool related measures. Both Ebert and Jurek et al. were unable to show symptom improvement, however, these studies were notably heterogeneous in their design. This is evident in their use of resonant frequency, as only one of them uses resonant frequency (Stern et al) while the other does not (Liu et al). When considering the two studies that did not show a significant improvement, these showed the same distribution, with Ebert utilising resonant frequency and Jurek et al not. It is also evident in the length of follow-up for the papers that stated them, as Liu et al which showed significant improvement in their results had a longer follow-up period of six weeks compared to the four weeks seen in Jurek et al’s trial. Due to small sample sizes (n= 24, 26 and 14, with the exception of Liu et al, n=85), the power of these studies (although not otherwise stated) would be relatively low. In summary, the accumulated evidence from the reviewed papers indicates that biofeedback could be useful as a potential therapy for FGIDs, but more investigation is required to further assess its efficacy. (**Table 2**)

Jurek et al stated the compliance of their participants to the SDB intervention, with six out of the seven participants completing at least 80% of their SDB sessions over the four weeks with an average of 19 sessions being completed (Katherine Jurek et al. 2022). Stern et al did not give a measure of compliance to the HRVB intervention but rather stated the number of sessions completed over the study period, which ranged from three to 19 sessions, however they stated that all participants who returned to follow-up experienced some benefit (Stern, Guiles, and Gevirtz 2014). Neither Ebert or Liu et al mentioned compliance to their study intervention.

Of the two studies that included a control group (Liu et al. and Jurek et al.) within their protocol, both found no significant changes in the IBS-SSS sores over the study, compared to baseline. One of the studies (Liu et al. 2022) found a trend that could be indicative of such a period of time to find efficacy. Within their study, they completed follow ups at weeks three and six after the commencement of the slow, deep breathing exercise (SDB). During these follow-ups, a trend emerged where many of the GI based outcomes measured, only started showing a difference in the SDB group compared to the sham group at the six week follow-up and not the three week follow-up. This trend was present for the IBS-SSS, BSFS, weekly complete spontaneous bowel motions and weekly spontaneous bowel motions. The same was found for the HRV metrics, keeping in trend with what would be predicted from a sham group (**Refer to Table 2**). Jurek et al also followed a similar trend with their study where they did not show a significant improvement in the recorded metrics at their four week follow-up mark. These same conclusions cannot be drawn for Stern et al and Ebert as neither of these studies stated their follow-up periods for their participants.

## Discussion

This scoping review has systematically evaluated the current literature regarding HRV Biofeedback and its potential use as a therapy for FGID, with a particular focus on the protocols and outcomes each study has employed. The studies identified had a heterogeneous design, with half being randomised controlled trials and the other being non-randomised interventional studies. Half of the studies identified showed that HRV Biofeedback had a beneficial effect on FGID symptoms, however, significant heterogeneity was identified. This review highlights the potential role for HRV biofeedback in FGIDs, while highlighting that duration of biofeedback training as a potential key parameter for treatment efficacy and providing guidance for advancing future studies based on the existing literature.

### Biofeedback Protocol and Metrics

All of the studies identified followed similar structures for their Biofeedback protocols, with only minor differences in methodology. All had participants trained to conduct biofeedback in the clinic/lab, and then had them continue to regularly practise biofeedback for up to six weeks. They also had all participants complete a questionnaire about their symptoms over their individual studies. The differences emerge when the breathing paces, HRV measurements, gastrointestinal symptom measures are considered.

#### Resonance Frequency

One key point of variation emerging from this review is whether study investigators instructed the participants to breathe at a standardised breathing frequency (i.e. 6 breaths per min) (Liu et al and Juek et al) or at the participants resonance frequency (Stern et al and Ebert). Resonance frequency is a theory that is often used when conducting biofeedback. It is the frequency of breathing where the oscillation of heart rate due to the respiratory sinus arrhythmia (produced by the slow breathing) resonates with the oscillation in heart rate due to the baroreflex. This results in a maximal heart rate variation, and a maximal increase in baroreflex gain with an increase in baroreflex gain at resting states after consistent practice as well (Vaschillo, Vaschillo, and Lehrer 2006; P. M. Lehrer et al. 2003; Fredric Shaffer 2020). The research into the benefit of employing resonance frequency into biofeedback training is limited, although an analogue study conducted in 2017 found that using a resonant frequency compared to a standardised breathing frequency was associated with a higher positive mood and a significantly higher LF/HF HRV ratio (Steffen et al. 2017). The method to find one’s resonant frequency typically involves trialling several different breathing frequencies for a short period of time and assessing the resultant LF power, HR_max_ - HR_min_, and participant comfort to find the optimum frequency (P. Lehrer et al. 2013; Fred Shaffer and Meehan 2020). Variations in resonant frequency can be influenced by one’s height and sex, with taller individuals and men having lower resonance frequencies than shorter individuals and women. However, most people tend to have resonant frequencies within a tight range of 5 to 6.5 breaths/min (Vaschillo, Vaschillo, and Lehrer 2006; P. Lehrer 2013). There is some evidence that one’s resonant frequency is not a stable metric, with one study finding a change in resonant frequency with 66.7% of its participants (Capdevila et al. 2021); however this was limited to a change in the mean of 0.2 breaths/min. When considering that most biofeedback systems are only able to adjust the breathing pacer in 0.5 breaths/min increments, this change in resonant frequency between tests may be clinically negligible. This data suggests that resonant frequency is a viable method to use to optimise biofeedback training, however the results of this scoping review indicate that more research is required into the topic to further understand the extent of its benefit as the two studies that showed significant results (Stern et al and Liu et al) and the two studies that did not (Ebert and Jurek et al), both had an equal distribution whether they used resonant frequency or a standardised breathing rate.

#### HRV Measurement and Analysis

All the studies employed the use of ECG in the clinic/lab setting to monitor the HRV of participants during the biofeedback exercises. ECG (often measured over 24 hours) is considered the gold standard of HRV measurement (Fred Shaffer and Ginsberg 2017). However, a question has emerged about the validity of other forms of HRV measurement. The main contender to the ECG is the photoplethysmography (PPG) method. This method relies on a light source emitting into the participant’s peripheral artery anywhere on the body (i.e. radial artery or digital artery), a proportion of this light is then absorbed according to the volume of blood in the artery at any one time, and the rest is reflected back towards the PPG device to be sensed as used in Stern et al. Because the volume of blood in the artery varies in accordance with the cardiac cycle, the PPG gives a reliable measure of the pulse rate and thus, by extension, the heart rate (Alian and Shelley 2014). This method is much more portable and accessible than ECG, with its key drawback being that the ECG allows for better theoretical detection of ectopic beats with its ability to show the electrical activity of the heart. However, a recent study shows physicians were able to detect atrial fibrillation using PPG measurement with equivalent accuracy to single-lead ECG (Gruwez et al. 2021), and therefore PPG may be feasible. PPG also has the added benefit of being conducted with a participant’s smartphone using its inbuilt flash and camera, therefore being highly accessible, and that this method, when combined with applications that employ the biofeedback principles, is able to detect HRV to accurately (Vandenberk et al. 2017; Plews et al. 2017).

In terms of how the studies chose to analyse their HRV data through their trials, all of them calculated a spectral analysis to display the power of the different frequency domains; three of them measured SDNN and RMSSD, and two of them calculated some form of stress index. There appears to be a lot of commonality between these studies in terms of how they measure, display and analyse HRV metrics. The spectral analysis of an individual’s HRV data via a Fast Fourier Transform provides useful information about one’s autonomic function, with changes in power in certain frequency bands being related to changes in sympathetic and parasympathetic activity (Fred Shaffer and Ginsberg 2017). This spectral analysis is often expressed as a ratio of LF/HF to analyse the balance of the sympathetic and parasympathetic systems. The assumption behind this is that LF power and HF power both correspond to sympathetic and parasympathetic activity respectively (Fred Shaffer, McCraty, and Zerr 2014; Pagani et al. 1984). However, this has been challenged in the past as the SNS and PNS are not solely influenced by LF and HF power. There is often some cross-over between them along with confounding due to baroreflex activity and respiration mechanics (Billman 2013). This is supported by the evidence that SDNN and RMSSD are commonly used metrics to describe the variation in heart rate with both of them being strongly correlated to autonomic activity and its influence on heart rate and respiratory sinus arrhythmia (Fred Shaffer, McCraty, and Zerr 2014; Fred Shaffer and Ginsberg 2017). Both of these are also greatly correlated to the spectral analysis of heart rate, with SDNN being associated with changes in ULF, VLF and LF power and RMSSD being highly correlated to HF power.

The SI (**Fig 2**) is another measure that is used as an analogue of sympathetic activity. First developed by Baevsky, this metric is highly sensitive to changes in sympathetic tone both within emotional and physical stress situations (Baevsky and Chernikova 2017). SI has been validated within its use in psychosomatic self-regulation, although evidence of its validation in biofeedback studies is scarce (Ognev et al. 2019).

In addition, although there are many time periods over which HRV is measured and these metrics can be calculated, the standard minimum period of time required to get a measurement of any of these is five minutes (Fred Shaffer and Ginsberg 2017).

#### Symptom Measurement

All four studies identified had similarities in having a strong focus on the symptoms associated with the specific FGID subtype they were investigating. Only one of the studies conducted a clinical test, specifically anorectal manometry, in order to identify the changes in the threshold of anorectal sensation (Liu et al. 2022). This focus on symptoms is likely due to the historic focus on the symptoms of FGID, such as in the Rome criteria of diagnosis, as relatively little is conclusively known about the physiology underlying these disorders (“Rome IV Criteria” 2020). The most common symptom scoring used here was the IBS-SSS (used by Liu et al and Jurek et al). This scale is well tested psychometrically and is easy to use with a good reproducibility. However, the main drawback of this tool is that it lacks adequate correlation with other abdominal pain measurement tools (Mujagic et al. 2015). Only one of the studies also recorded the impact of these symptoms on the participant’s quality of life (Stern, Guiles, and Gevirtz 2014), an impact of these disorders which can sometimes be overlooked by clinicians (Rocque and Leanza 2015). Due to the lack of physiological biomarkers within FGID, there are new methods being developed, giving researchers and clinicians an insight into the physiology of FGIDs and providing biomarkers for analysis. One such example is Body Surface Gastric Mapping (BSGM), a method that can measure gastric electrical activity with high accuracy and correlation to symptomatology, providing a new understanding of the physiological basis of FGID symptoms (Schamberg et al. 2023). This is also a potential tool that can be used to provide an objective measurement and understanding of the symptoms that participants experience in future trials, using a validated app-based approach (Sebaratnam et al. 2022).

It is clear that greater standardisation within the HRVB protocol is needed, with the variables chosen being based on the current evidence of what increases HRV and autonomic flexibility.

### Interventional and Control Group Results

Out of the four studies that were conducted, only two found a statistically significant improvement in symptoms for the group of participants who partook in the HRV biofeedback intervention (Stern, Guiles, and Gevirtz 2014; Liu et al. 2022). Out of the four studies, only two of them employed the use of a control group as a comparison for the biofeedback groups results (Liu et al. 2022; Jurek et al. 2022), while the other two did not employ a control or similar method (Stern, Guiles, and Gevirtz 2014; Ebert 2012). The studies that did use a control group found no statistically significant difference in any measurement.

Only two of the studies found a significant improvement in symptoms after partaking in biofeedback. This is likely due to the heterogeneity of the protocols exemplified earlier in the review. Only two of the studies employed the use of resonance frequency compared to a standardised breathing frequency, and the duration of the biofeedback intervention varies between each study, along with the time that is spent practising biofeedback while at home. One key finding that was found during this review was that it is likely that biofeedback will need to be consistently practised for at least six weeks for its effects to become evident. This finding was demonstrated when examining both Jurek et al’s and Liu et al’s trials. Jurek et al only had their participants practise biofeedback for four weeks while Liu et al had their participants practise SDB for six weeks. Where Jurek et al did not find any significant improvements in symptoms, Liu et al did. And upon closer inspection into Liu et al’s findings, these differences only started to become significant after 4 weeks into Liu et al’s trial. Thus, it is possible that both consistency and duration of biofeedback training is an important factor predictive of improvement of clinical FGID symptoms. However, the evidence behind this theory is drawn from only two trials and thus more evidence is needed to support it. The remaining two studies (Ebert and Stern et al) do not state how long their follow up periods are and so we are not able to draw this conclusion from them.

### Limitations of this Review

There are several limitations to this review, including its focus on FGID without considering the potential for other disorders both within and outside the gastrointestinal system, such as within the urinary system (Zivkovic et al. 2017). This review does not investigate the effect of these biofeedback interventions on the participants HRV metrics over their respective trials due to its focus on the change in FGID symptoms. This review also did not assess the variance in baseline HRV for those diagnosed with FGID compared to healthy controls as other reviews have done similar feats (Ali et al. 2023). Finally, we were not able to conduct a quantitative analysis of the studies identified due to their small sample sizes and heterogeneity in participant population, FGID subtype, and biofeedback protocol.

### Future Research

This review demonstrates that the field of HRVB is promising yet still in its infancy and thus more research into the impact of HRVB on FGID is needed, particularly more randomised controlled trials to better assess the effect of biofeedback compared to treatment as usual or no treatment.

Future studies should also continue to evaluate the use of PPG as a method to measure HRV compared to a single lead ECG, especially when participants are outside of the clinic or lab. PPG has a high level of accessibility and has an ability to be used alongside a smartphone application for biofeedback exercises. And although the benefit of ectopic beat detection is reduced with PPG compared to ECG, the benefit of accessibility could far outweigh the limitations of artefact removal. This makes it a tool that can potentially improve the way that biofeedback is conducted in clinical trials and opens the door to assess how HRV metrics change with each session completed with an almost similar accuracy compared to ECG.

The utilisation of more clinical tools can further assess the underlying physiology beyond just the symptoms of FGIDs. This could result in a more objective measurement of how an individual’s gastrointestinal physiology changes during the biofeedback intervention, thus allowing for greater advancements in FGID diagnosis and treatment options.

## Data Availability

All data produced in the present study are available upon reasonable request to the authors

